# DNA Methylation Biomarkers Capture Residual Biological Risk Beyond PREVENT

**DOI:** 10.64898/2026.08.07.26359993

**Authors:** Diensn G. Xing, Md. Shenuarin Bhuiyan, Steven A. Conrad, Arif Yurdagul, Oren Rom, A. Wayne Orr, Christopher G. Kevil, SM Ashiqul Islam, Mohammad Alfrad Nobel Bhuiyan

**Affiliations:** Department of Biochemistry, LSU Health Shreveport; Department of Medicine, Louisiana State University Health Sciences Center at Shreveport, Shreveport, LA, 71103, USA; Department of Pathology and Translational Pathobiology, Louisiana State University Health Sciences Center at Shreveport, Shreveport, LA, 71103, USA; Department of Molecular and Cellular Physiology, Louisiana State University Health Sciences Center at Shreveport, Shreveport, LA, 71103, USA; Department of Epidemiology and Biostatistics, College of Integrated Health Sciences, University at Albany– State University of New York, Albany, NY

**Keywords:** DNA methylation, epigenetic clocks, mortality, risk assessment, cardiovascular disease, PREVENT

## Abstract

**Background:** Contemporary cardiovascular disease (CVD) risk equations may not fully capture cumulative biological aging or long-term exposure burden. DNA methylation (DNAm) biomarkers may capture aging- and exposure-related biology, but their incremental prognostic value beyond clinical risk-factor models like PREVENT remains uncertain. To our knowledge, no prior study has benchmarked DNAm-based biomarkers with PREVENT.

**Methods:** In a population-based cohort study, we analyzed NHANES 1999-2002 participants with DNAm biomarkers and mortality follow-up. We derived a DNAmScore from candidate DNAm biomarkers using elastic-net Cox regression with repeated nested cross-validation. A PREVENT-like clinical model was defined as a Cox model fit in NHANES using PREVENT predictors. Weighted Cox models estimated the association between DNAmScore and mortality after adjustment for PREVENT-like clinical predictors. We then compared the PREVENT-like clinical model, DNAmScore alone, and a combined model (PREVENT-like clinical predictors plus DNAmScore) using cross-fitted C-index, time-dependent AUC, calibration, and Brier score.

**Results:** Our cohort included 2,282 participants; 597 and 937 deaths occurred by 10 and 15 years, respectively. After adjustment for PREVENT-like clinical predictors, the cross-fitted DNAmScore was strongly associated with all-cause mortality (HR per 1-SD increase, 2.43; 95% CI, 1.97–2.99). At 10 years, AUCs were 0.791 for the PREVENT-like model, 0.791 for DNAmScore, and 0.803 for the combined model. At 15 years, corresponding AUCs were 0.825, 0.822, and 0.835. Compared with the PREVENT-like model, the combined model improved AUC by 0.013 (95% CI, 0.006–0.020) at 10 years and 0.010 (95% CI, 0.004–0.015) at 15 years. The combined model had lower Brier scores at all three horizons with similar calibration. DNAmScore remained associated with CVD mortality after clinical adjustment.

**Conclusions:** DNAmScore identified residual biological risk beyond PREVENT-like clinical predictors, with strong independent mortality associations and modest, consistent improvements in cross-fitted prediction performance. These findings support development and external validation of CVD-specific DNAm biomarkers.

**What Is Known:**

- Contemporary cardiovascular risk models may not fully capture cumulative biological aging and exposure burden.
- DNA methylation biomarkers are associated with mortality, but their incremental predictive value beyond clinical risk predictors remains uncertain.

**What the Study Adds:**

- In a nationally representative NHANES cohort, a cross-fitted composite DNAmScore remained strongly associated with mortality after adjustment for PREVENT predictors.
- Adding DNAmScore produced modest improvements in discrimination and prediction error at 5, 10, and 15 years, and DNAmScore was also associated with cardiovascular mortality in secondary analyses.

## Introduction

Cardiovascular disease (CVD) is the leading cause of death globally and in the US^1^. Many forms of CVD, including atherosclerotic and cardiometabolic disease, develop over decades, making early prevention and risk estimation particularly important, with risk equations playing a direct role in guiding shared decision making^2^. Currently, CVD risk estimation increasingly incorporates the PREVENT equations^2,3^, which are based on traditional cardiometabolic risk factors^4^. However, these models measure risk factors at a single visit and may not fully capture cumulative biological aging or exposure burden.

A recent focus of biomarker and aging research is the development of DNA methylation (DNAm)-based aging measures (i.e., epigenetic clocks)^5,6^. Early epigenetic clocks were trained to estimate chronological age, whereas later-generation clocks were trained on phenotypic aging, lifespan, mortality, or pace-of-aging^5,7,8^. These newer clocks have shown stronger associations with mortality and age-related disease than those based on chronological age, and may provide additional risk information beyond chronological age alone^6^. Because DNAm patterns are associated with aging and can reflect environmental, lifestyle, and health-related exposures (e.g., smoking and metabolic risk factors), DNAm-based measures may serve as integrative markers of cumulative biological risk^9–11^. This is particularly relevant to CVD, a disease in which long-term lifestyle and behavioral exposures contribute substantially to risk^12–14^.

Despite growing evidence demonstrating the associations between epigenetic clocks and mortality^15^, it is unclear whether they provide incremental risk information beyond clinical models alone. This is critical because clinical predictors and DNAm biomarkers may capture complementary aspects of risk: clinical models measure current cardiometabolic burden, whereas DNAm clocks may reflect cumulative aging and exposures. Furthermore, prior studies have focused on individual clocks or age-acceleration measures rather than deriving a DNAm score from multiple candidates. In this study, we use penalized survival modeling to derive a DNAmScore from candidate DNAm biomarkers, then evaluate its independent association with mortality and incremental predictive value compared with PREVENT.

## Methods

### Study Population and Data Sources

We analyzed participants from the 1999–2000 and 2001–2002 cycles of the National Health and Nutrition Examination Survey (NHANES), a nationally representative survey of the noninstitutionalized United States population^16^. Publicly available NHANES demographic, examination, laboratory, dietary, and questionnaire files were merged by participant sequence number. DNA methylation (DNAm) biomarker data were linked to the NHANES analytic file using participant identifiers. Participants were eligible if they had measured DNAm biomarkers, a positive DNAm subsample weight, linked mortality follow-up, nonmissing follow-up time, and complete data for the clinical predictor set^17^. No imputation was used.

Of 21,004 NHANES 1999–2002 participants in the merged source dataset, 2,532 had a positive 4-year DNAm subsample weight, measured DNAm biomarkers, and valid linked mortality follow-up. Among these participants, 231 were excluded because at least one PREVENT-like clinical predictor was incomplete, leaving 2,301 participants. An additional 19 participants were excluded because required DNAm predictor values were unavailable, leaving a final complete-case analytic cohort of 2,282 participants. During follow-up, 1,200 participants died, including 252 deaths by 5 years, 597 by 10 years, and 937 by 15 years. No imputation was performed (Figure 1)

**Figure 1:**
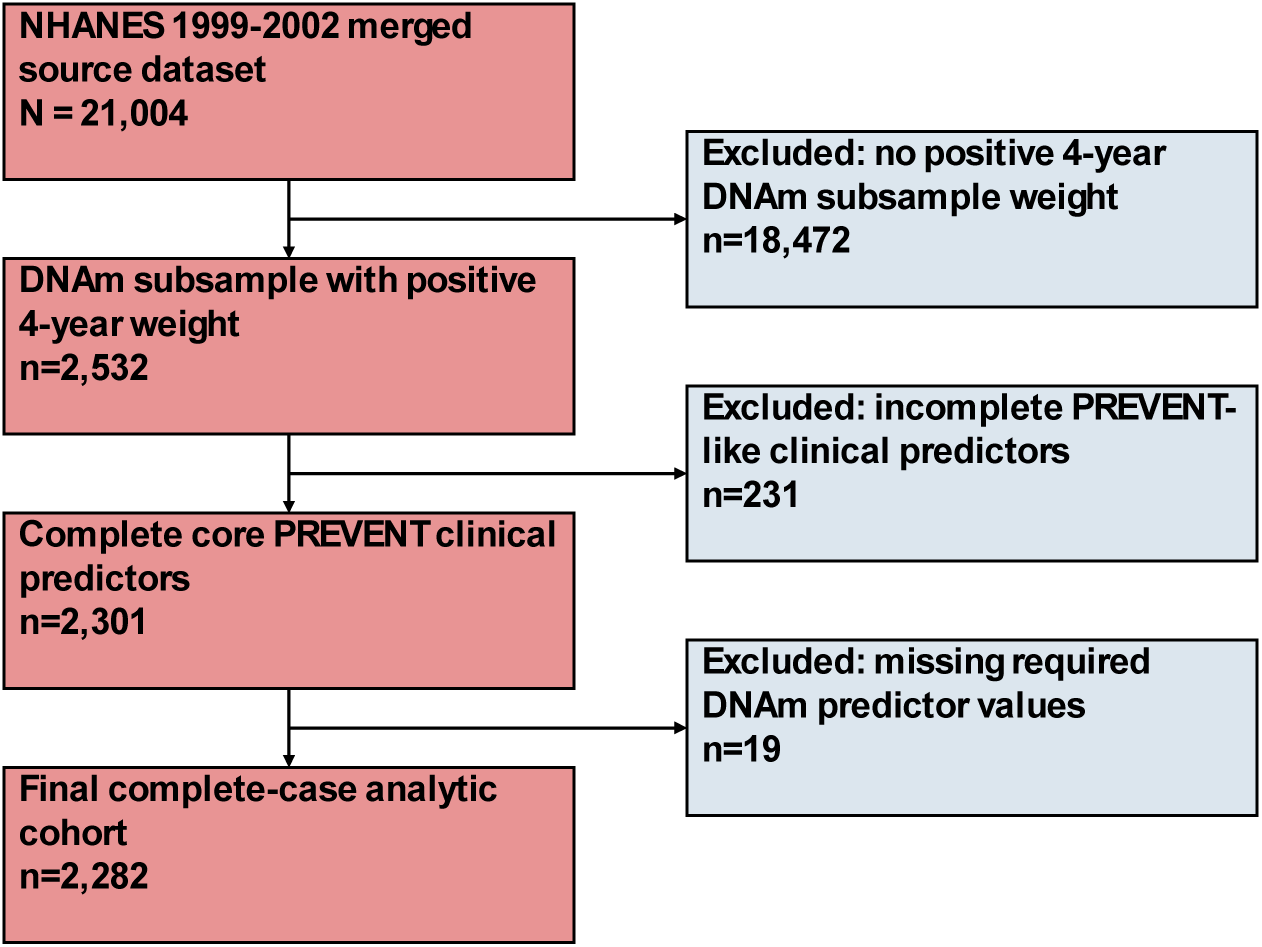
Cohort Inclusion Criteria. The final analytic cohort required correct DNAm subsample weights, complete PREVENT-like clinical predictors, and complete DNAm biomarker values.

### Mortality Outcomes

Mortality status and follow-up time were obtained from the 2019 NHANES public-use linked mortality files, with follow-up through December 31, 2019^16^. The primary outcome was all-cause mortality. Follow-up time was calculated using the available months from examination or interview to death or censoring. For secondary analyses, cardiovascular disease mortality was defined using underlying cause-of-death codes from the linked mortality files, with non-CVD deaths treated as competing events in competing-risk models.

### Clinical Predictor Set

To compare DNAmScore with PREVENT as closely as possible, we refit a Cox model within NHANES ^4^ using the available PREVENT predictor set. The published PREVENT coefficients were not directly applied because those equations were developed for incident CVD outcomes rather than all-cause mortality; refitting also allowed all three models to be evaluated for the same outcome within the same internal-validation framework. Specifically, we fit a multivariable Cox model in our cohort with age, sex, body mass index, systolic blood pressure, total cholesterol, high-density lipoprotein cholesterol, estimated glomerular filtration rate, diabetes, current smoking, antihypertensive medication use, and lipid-lowering medication use as covariates. Estimated glomerular filtration rate was calculated using the 2021 race-free CKD-EPI creatinine equation. Diabetes was defined using self-reported physician diagnosis or laboratory criteria, including hemoglobin A1c ≥6.5% or fasting plasma glucose ≥126 mg/dL. Current smoking was defined using self-reported current smoking status, with serum cotinine ≥10 ng/mL used when questionnaire-based smoking status was missing. Anti-hypertensive and lipid-lowering medication variables were derived using NHANES blood pressure and cholesterol questionnaires using skip-pattern logic^18^.

### DNA Methylation Biomarkers

Candidate DNAm predictors included established epigenetic clocks, DNAm mortality-related biomarkers, and estimated blood-cell proportions. Epigenetic clocks included HorvathAge^5^, HannumAge^19^, SkinBloodAge^20^, PhenoAge^21^, GrimAgeMort^8^, GrimAge2Mort^7^, DunedinPoAm^22^, ZhangAge^23^, LinAge^24^, WeidnerAge^25^, and VidalBraloAge^26^. DNAm mortality-related biomarkers included DNAm surrogates for GDF15Mort, B2MMort, CystatinCMort, TIMP1Mort, ADMMort, PAI1Mort, LeptinMort, PACKYRSMort, CRPMort, and A1CMort^7,8,18^. Positively skewed biomarkers were log-transformed before modeling when appropriate. Twenty-seven candidate predictors were initially considered. Before penalized modeling, GrimAge2Mort was removed because of its high correlation with GrimAgeMort (*r*=0.986), and ZhangAge was removed because of its high correlation with SkinBloodAge (*r*=0.968). This left 25 predictors, with all retained pairs having absolute correlations <0.95.

### DNAmScore Derivation

DNAmScore was derived using unweighted elastic-net Cox regression^27,28^ with 25 repeats of nested 5-fold cross-validation, using 5 inner folds within each outer training set^29^. Candidate DNAm predictors were standardized within each outer training set. Values of α from 0.50 to 1.00 and candidate values of λ were jointly evaluated in the inner folds using a one-standard-error rule favoring the sparsest non-null model. The selected model was then applied to the held-out outer fold. Held-out scores were averaged across the 25 repeats and standardized using the DNAm subsample weights. The clinical, DNAmScore-only, and combined models were evaluated using the same outer folds. A separate all-data fit was used only to describe the biomarkers retained when the model was fit to the full analytic cohort.

### Statistical Analysis

Participants were summarized overall and by mortality status. Continuous variables were summarized using means and standard deviations or medians and interquartile ranges, as appropriate, and categorical variables were summarized as counts and percentages.

The association between cross-fitted DNAmScore and all-cause mortality was estimated using survey-weighted Cox regression with the DNAm subsample weights, masked strata, and masked primary sampling units^16,18^. DNAmScore hazard ratios were reported per 1-SD increase. The proportional-hazards assumption was evaluated using Grambsch–Therneau tests based on scaled Schoenfeld residuals. Because time-transform terms could not be fitted directly with the full survey-design model, extended Cox sensitivity models used normalized DNAm subsample weights and robust standard errors clustered by the combination of survey stratum and primary sampling unit ^30^.

Prediction analyses used out-of-fold predictions from repeated nested cross-validation. Discrimination was assessed using Harrell’s C-index and IPCW dynamic time-dependent AUC at 5, 10, and 15 years. Paired AUC differences were tested using the iid-representation method implemented in timeROC^31^, with *P* values corrected for the 3 horizon-specific comparisons within each model contrast. Calibration was assessed by comparing predicted risk with Kaplan– Meier observed risk across deciles, and prediction error was assessed using horizon-specific IPCW Brier scores^32,33^.

Subgroup analyses evaluated model performance across clinically relevant strata, including age, sex, smoking status, diabetes status, and race/ethnicity where event counts permitted.

For secondary CVD mortality analyses, survey-weighted cause-specific Cox regression was the principal sensitivity analysis, with non-CVD deaths censored at the time of death. Fine–Gray models treating non-CVD death as a competing event were performed as supplemental unweighted analyses. Both analyses reused the cross-fitted all-cause DNAmScore rather than deriving a CVD-specific score.

Analyses were performed in R^34^. Elastic-net Cox models were fit using glmnet^27^, including its Cox model implementation. Unweighted Cox models were fit using survival^30^, and survey-weighted Cox models were fit using survey^35^. Fine-Gray models, absolute-risk prediction, and prediction-error estimates were obtained using riskRegression^32^ and prodlim^33^. Time-dependent AUCs were estimated using timeROC^31^. Tables and figures were generated using tableone, gtsummary, ggplot2, and related tidyverse packages^36^.

## Results

### Analytic Cohort and Event Count

After filtering for all eligible complete cases, the analytic cohort included 2,282 NHANES 1999– 2002 participants with measured DNAm biomarkers, positive DNAm subsample weights, linked mortality follow-up, and complete PREVENT-like clinical predictors. There were 1,200 deaths during follow-up, including 252 by 5 years, 597 by 10 years, and 937 by 15 years. Participants who died were older, had higher systolic blood pressure and lower eGFR, and were more likely to be male, currently smoking, and diabetic. Total cholesterol was modestly lower among participants who died, whereas BMI did not differ significantly between groups **(Table 1)**.

**Table 1.** Baseline prediction cohort characteristics by mortality status. NHANES 1999-2002 prediction cohort, stratified by mortality status. Values are unweighted mean (SD) for continuous variables and n (%) for categorical variables. *p-*values compare alive vs deceased groups. DNAm = DNA methylation; eGFR = estimated glomerular filtration rate; HDL = high-density lipoprotein.

| Characteristic | Overall | Alive | Deceased | *P* value |
| --- | --- | --- | --- | --- |
| *N* | 2,282 | 1,082 | 1,200 | — |
| Age, years | 65.87 (9.86) | 60.46 (7.25) | 70.74 (9.35) | <0.001 |
| BMI, kg/m<sup>2</sup> | 28.59 (5.69) | 28.83 (5.44) | 28.37 (5.91) | 0.054 |
| Systolic BP, mmHg | 137.69 (22.34) | 132.55 (20.11) | 142.33 (23.24) | <0.001 |
| Total cholesterol, mg/dL | 210.75 (39.88) | 212.71 (38.23) | 208.99 (41.25) | 0.026 |
| HDL, mg/dL | 51.98 (16.05) | 52.23 (15.57) | 51.76 (16.48) | 0.486 |
| eGFR, mL/min/1.73 m<sup>2</sup> | 86.02 (20.52) | 92.36 (16.30) | 80.30 (22.19) | <0.001 |
| Male | 1,169 (51.2%) | 512 (47.3%) | 657 (54.8%) | <0.001 |
| Female | 1,113 (48.8%) | 570 (52.7%) | 543 (45.2%) | — |
| Nonsmoker | 1,896 (83.1%) | 927 (85.7%) | 969 (80.8%) | 0.002 |
| Current smoker | 386 (16.9%) | 155 (14.3%) | 231 (19.2%) | — |
| No diabetes | 1,730 (75.8%) | 875 (80.9%) | 855 (71.2%) | <0.001 |
| Diabetes | 552 (24.2%) | 207 (19.1%) | 345 (28.7%) | — |
| Mexican American | 658 (28.8%) | 348 (32.2%) | 310 (25.8%) | <0.001 |
| Other Hispanic | 141 (6.2%) | 85 (7.9%) | 56 (4.7%) | — |
| Non-Hispanic White | 947 (41.5%) | 400 (37.0%) | 547 (45.6%) | — |
| Non-Hispanic Black | 463 (20.3%) | 201 (18.6%) | 262 (21.8%) | — |
| Other/multiracial | 73 (3.2%) | 48 (4.4%) | 25 (2.1%) | — |

### DNAmScore Derivation

After correlation pruning, DNAmScore was derived from 25 candidate predictors. Joint inner cross-validation selected α=1.00 in 124 of 125 outer training models and α=0.90 in 1 model. Outer models retained a mean of 4.62 biomarkers. GrimAgeMort, TIMP1Mort, B2MMort, and HannumAge were selected in 100%, 95.2%, 88.8%, and 87.2% of outer models, respectively; PhenoAge and HorvathAge were selected in 47.2% and 32.8%. The separate all-data model, reported descriptively and not used for performance estimation, selected α=1.00 and λ=0.0415 and retained GrimAgeMort, HannumAge, TIMP1Mort, B2MMort, HorvathAge, CRPMort, and PhenoAge. Survival differed across DNAmScore tertiles (*p*<0.001) **(Supplemental Figure S1)**.

### DNAmScore and Association with Mortality

In the survey-weighted Cox model, higher cross-fitted DNAmScore was strongly associated with all-cause mortality after adjustment for PREVENT clinical predictors. Each 1-SD increase in DNAmScore was associated with a 2.43-fold higher mortality hazard (HR, 2.43; 95% CI, 1.97– 2.99; P<0.001). Diabetes (HR, 1.52; 95% CI, 1.28–1.82), older age (HR per 10 years, 1.28; 95% CI, 1.02–1.59), higher systolic blood pressure (HR per 10 mmHg, 1.06; 95% CI, 1.02–1.10), and female sex (HR versus male sex, 0.74; 95% CI, 0.61–0.91) showed the strongest clinical associations **(Figure 2)**. The global proportional-hazards test was significant, but only lipid-lowering medication was significant in term-specific testing (*P*=0.041). DNAmScore showed no evidence of nonproportionality in either the Schoenfeld-residual test (*P*=0.529) or the DNAmScore-by-log-time test (*P*=0.302). Allowing age and lipid-medication effects to vary with time produced nearly identical DNAmScore estimates **(Supplemental Table S1)**.

**Figure 2:**
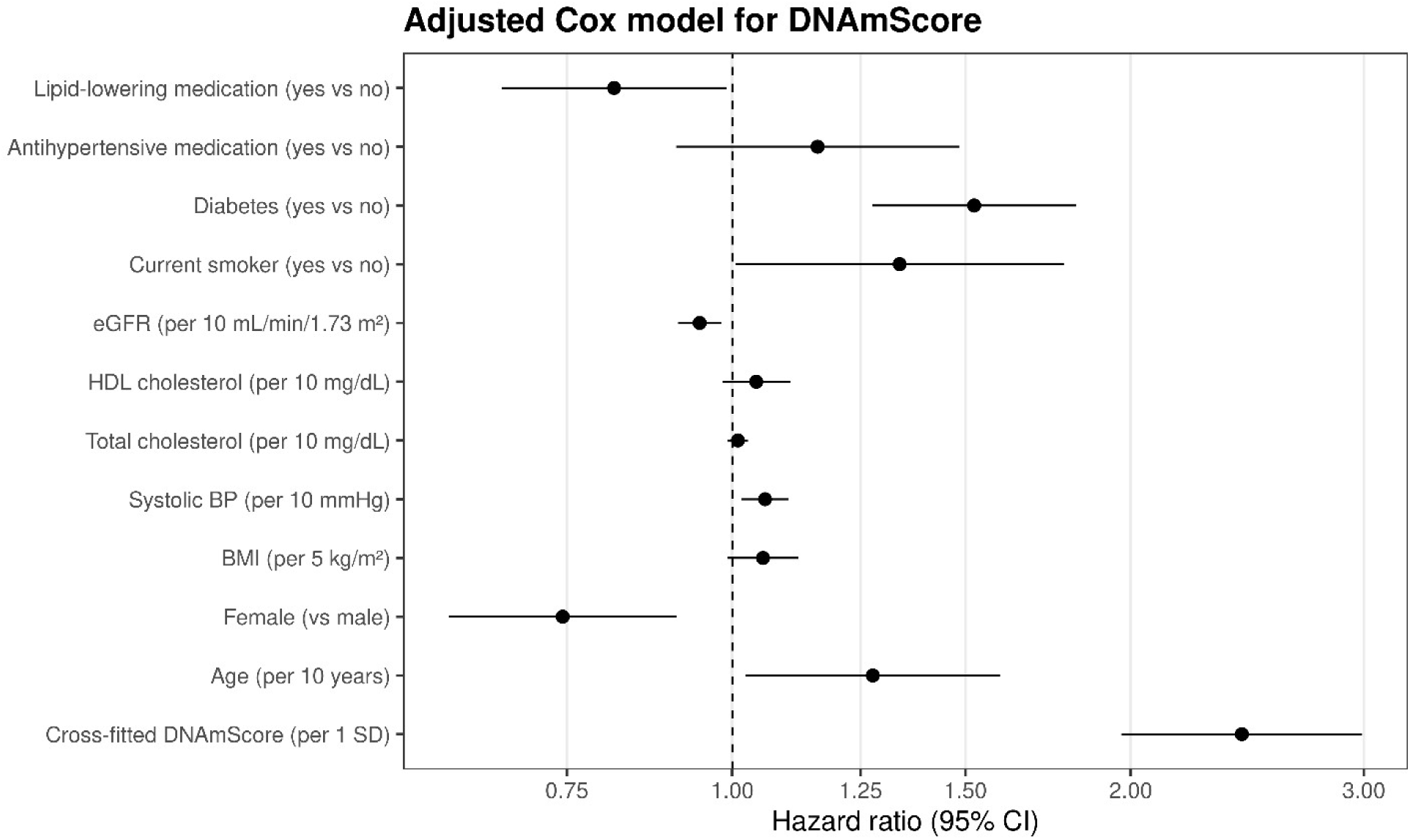
Adjusted hazard ratios from the survey-weighted robust Cox model including PREVENT-like clinical predictors and cross-fitted DNAmScore. Points indicate HRs and horizontal bars indicate 95% CIs on a log scale (HR=1 shown by dashed line). DNAmScore remains strongly associated with all-cause mortality after clinical adjustment.

### Discrimination

The combined model had the highest cross-fitted discrimination across all 3 horizons. Mean C-index values were 0.756 for PREVENT, 0.754 for DNAmScore, and 0.766 for the combined model. At 5 years, AUCs were 0.766, 0.772, and 0.784, respectively. Corresponding AUCs were 0.791, 0.791, and 0.803 at 10 years and 0.825, 0.822, and 0.835 at 15 years. Compared with PREVENT, adding DNAmScore improved AUC by +0.018 (95% CI, 0.008–0.028) at 5 years, +0.013 (95% CI, 0.006–0.020) at 10 years, and +0.010 (95% CI, 0.004–0.015) at 15 years. The combined model exceeded PREVENT at all horizons (*P*≤0.004). DNAmScore alone did not differ significantly from PREVENT at any horizon **(Figure 3 and Table 2)**.

**Figure 3:**
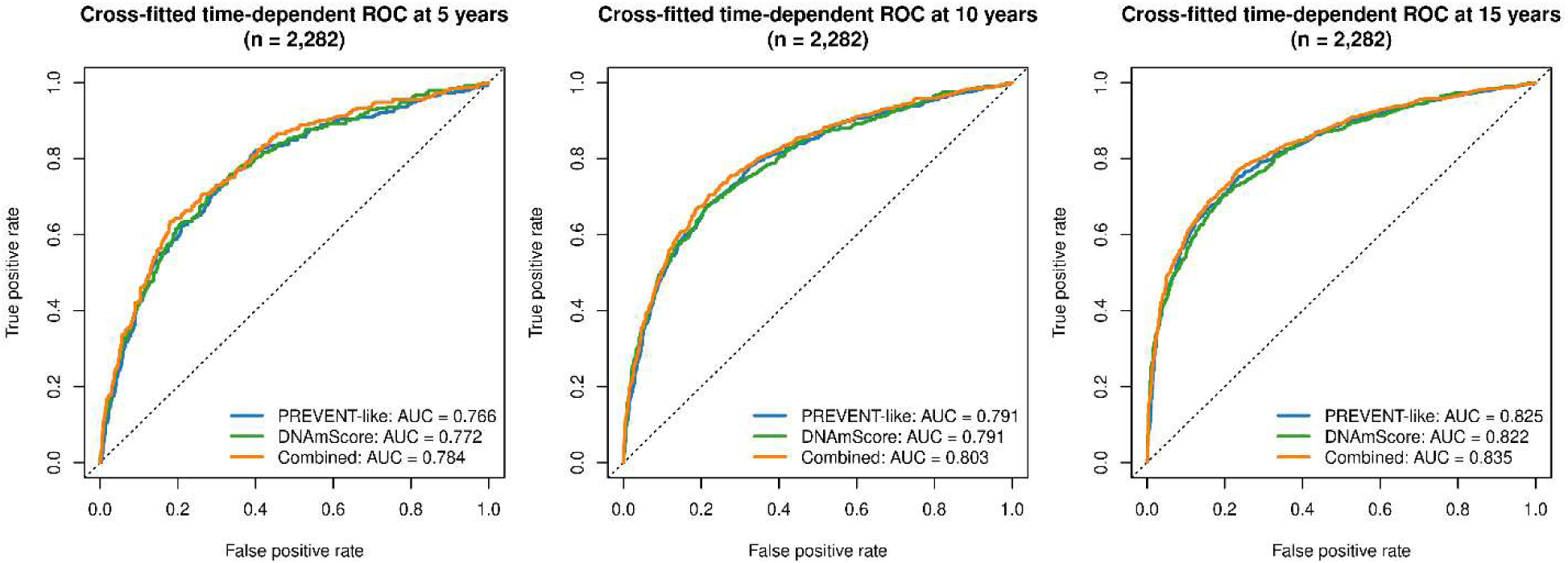
ROC curves at 5, 10, and 15 years comparing discrimination for the PREVENT-like model, DNAmScore alone, and the combined model (n=2282).

**Table 2.** Cross-fitted discrimination of PREVENT-like, DNAmScore, and combined models for all-cause mortality.

| **Metric** | **PREVENT** | **DNAmScore** | **Combined** | **ΔCombined versus PREVENT (95% CI)** | **P-value** |
| --- | --- | --- | --- | --- | --- |
| C-index | 0.756 (0.012) | 0.754 (0.013) | 0.766 (0.012) | +0.009 | — |
| 5-year AUC | 0.766 (0.734–0.798) | 0.772 (0.740–0.803) | 0.784 (0.754–0.815) | +0.018 (0.008–0.028) | <0.001 |
| 10-year AUC | 0.791 (0.769–0.813) | 0.791 (0.769–0.813) | 0.803 (0.782–0.825) | +0.013 (0.006–0.020) | 0.001 |
| 15-year AUC | 0.825 (0.808–0.843) | 0.822 (0.804–0.840) | 0.835 (0.818–0.852) | +0.010 (0.004–0.015) | 0.004 |

*Note: C-index values are mean (SD) across 125 outer models. AUCs are cross-fitted IPCW time-dependent AUCs with pointwise 95% CIs conditional on the fitted out-of-fold predictions. ΔAUC confidence intervals are paired and pointwise. P-values were corrected for the 3 horizon-specific comparisons within the combined-PREVENT contrast.*

### Calibration

Descriptive cross-fitted calibration was similar across the 3 models. For the combined model, mean predicted and observed risks were 0.111 and 0.110 at 5 years, 0.262 and 0.262 at 10 years, and 0.412 and 0.411 at 15 years. Combined-model calibration slopes were 0.934, 0.949, and 1.010 at 5, 10, and 15 years, respectively. IPCW Brier scores were lower for the combined model than for PREVENT at 5 years (0.0852 versus 0.0880), 10 years (0.1436 versus 0.1482), and 15 years (0.1577 versus 0.1627) **(Table 3 and Supplemental Figures S2 and S3)**.

**Table 3.** Cross-fitted calibration and IPCW Brier scores.

| Horizon | Model | Mean predicted risk | Observed risk | Intercept | Slope | IPCW Brier score |
| --- | --- | --- | --- | --- | --- | --- |
| 5 y | PREVENT | 0.110 | 0.110 | -0.155 | 0.906 | 0.0880 |
| 5 y | DNAmScore | 0.110 | 0.110 | -0.103 | 0.938 | 0.0865 |
| 5 y | Combined | 0.111 | 0.110 | -0.113 | 0.934 | 0.0852 |
| 10 y | PREVENT | 0.260 | 0.262 | -0.021 | 0.962 | 0.1482 |
| 10 y | DNAmScore | 0.262 | 0.262 | -0.031 | 0.969 | 0.1468 |
| 10 y | Combined | 0.262 | 0.262 | -0.045 | 0.949 | 0.1436 |
| 15 y | PREVENT | 0.412 | 0.411 | 0.006 | 1.034 | 0.1627 |
| 15 y | DNAmScore | 0.412 | 0.411 | 0.004 | 1.036 | 0.1644 |
| 15 y | Combined | 0.412 | 0.411 | -0.005 | 1.010 | 0.1577 |

*Note: Lower Brier scores indicate lower prediction error. Calibration slopes near 1.0 and* *intercepts near 0 indicate closer agreement between predicted and observed risk.*

### CVD Mortality

In the survey-weighted cause-specific Cox analysis, each 1-SD increase in cross-fitted DNAmScore was associated with CVD mortality before adjustment (HR, 3.70; 95% CI, 3.23– 4.23) and after adjustment for PREVENT clinical predictors (HR, 2.19; 95% CI, 1.62–2.96; *P*<0.001). In the supplemental unweighted Fine–Gray analysis, the corresponding adjusted subdistribution HR was 1.39 (95% CI, 1.09–1.77; *P*=0.007) **(Supplemental Table S2)**.

### Subgroup Analysis

The combined model generally produced higher cross-fitted AUCs than PREVENT across exploratory subgroups, although the magnitude of improvement varied. Among participants younger than 65 years, ΔAUC was +0.028 at 10 years and +0.018 at 15 years; corresponding differences among participants aged 65 years or older were +0.016 and +0.016. Point estimates were lower for the combined model among Mexican American participants at both horizons and among Other Hispanic participants at 15 years **(Supplemental Table S3 and Supplemental Figure S4)**.

## Discussion

In our study, we found that DNAm-based biomarkers were significantly associated with mortality, even after adjustment for PREVENT-like predictors^4^. Furthermore, adding DNAmScore to a PREVENT-like model produced small improvements in discrimination and prediction error, with similar calibration. In secondary analyses, DNAmScore was also associated with CVD mortality after adjustment for PREVENT-like predictors. Together, these findings support our hypothesis that DNAm-based biomarkers capture residual biological risk beyond traditional clinical predictors alone.

A key contribution of this study is the evaluation of DNAm biomarkers compared to a standard clinical risk model. Prior studies have shown that epigenetic clocks, particularly later-generation clocks such as GrimAge and GrimAge2, are associated with all-cause and cause-specific mortality^7,8^. However, many prior analyses have focused on individual clocks, epigenetic age acceleration measures, or association models adjusted for selected covariates. To our knowledge, few studies have evaluated whether DNAm epigenetic clocks and DNAm mortality biomarkers improve mortality prediction beyond PREVENT. Therefore, our analysis extends the existing epigenetic-clock literature from association testing toward clinical prediction.

The strong cross-fitted association with mortality after adjustment (HR, 2.43) supports our hypothesis that DNAmScore captures exposure-related biology not fully represented by one-time clinical measurements. PREVENT predictors (such as BP, cholesterol, diabetes, kidney function, current smoking, and medication use) mostly characterize current clinical burden^4^. DNAm-based biomarkers reflect cumulative smoking exposure and systemic inflammation, metabolic dysfunction, and aging-related biology^8,10^. Because these elements play an important role in CVD risk^12–14^, DNAm-based biomarkers that reflect longer-term exposure to these factors may indeed improve risk prediction.

Despite the strong association with mortality, Brier score and discrimination were only modestly better in the combined model than in the PREVENT-like model alone, with similar calibration. This indicates that, though DNAm-based biomarkers may capture some residual biology,

PREVENT encompasses a significant amount of important prognostic information. However, NHANES is a limited cohort, and the modest improvement supports further development of CVD-focused DNAm biomarkers and testing in more diverse cohorts. In addition, greater AUC improvements were noted in those <65 years of age compared to those ≥65 years of age. Because participants with DNAm data were older adults, traditional clinical risk factors may have already accumulated enough prognostic information to provide accurate risk prediction^3^. DNAm biomarkers may be more informative earlier in life, before traditional cardiac risk factors^37,38^ fully develop. Future studies in younger and midlife cohorts should test whether DNAm-based biomarkers identify those at high biological risk despite low or intermediate clinical risk. This could support more informed risk stratification and earlier prevention.

Our findings also highlight the need for CVD-specific risk prediction. The DNAmScore in this study was derived from biomarkers designed for all-cause mortality and selected biomarkers that likely reflect broad aging, immune, inflammatory, and multimorbidity-related risk^7,8,15^. Thus, it is expected that the performance and associations would be strongest for all-cause mortality rather than CVD-specific outcomes. Future work may focus on developing CVD-specific DNAm scores, rather than relying on mortality clocks.

The composition of DNAmScore may help explain its performance. GrimAge^8^ was selected in all 125 outer models and was the dominant term in the final all-data fit. The TIMP DNAm surrogate^8^, beta-2-microglobulin DNAm surrogate^8^, and HannumAge^19^ were also selected in most outer models, while the final fit additionally retained HorvathAge^5^, the CRP DNAm surrogate^7^, and PhenoAge^21^. Collectively, the selected markers encompass systemic mortality-related biology, such as smoking, inflammation, immune, metabolic, and renal-related risk, rather than only focusing on chronological aging^5,7,8,19,21^. Importantly, the association between DNAmScore and CVD mortality remained statistically significant in the CVD mortality analysis, including after adjustment for PREVENT-like clinical variables. This supports future work developing CVD-specific risk scores.

This study has several strengths. By incorporating the NHANES linked mortality files, we were able to extend a cross-sectional survey design into a population-based longitudinal mortality analysis with long-term follow-up. Rather than evaluating a single clock or biomarker, we derived a DNAmScore from multiple candidates using penalized survival modeling. Prediction performance was evaluated using repeated nested cross-validation and out-of-fold predictions, reducing optimism from in-sample model fitting. Additionally, fitting the PREVENT-like clinical model in our cohort using the same cross-validation framework allowed an internally consistent comparison of DNAmScore and PREVENT. We also evaluated multiple domains of model performance, including discrimination, calibration, and prediction error, expanding beyond association alone, and we performed secondary CVD mortality analyses using both cause-specific Cox and Fine-Gray competing-risk models.

A few limitations should be noted. PREVENT was approximated using available NHANES variables rather than applying the formal published PREVENT equations, as noted above. DNAm and clinical predictors were measured at a single baseline examination. Publicly available longitudinal outcome data in NHANES are limited to linked mortality, preventing evaluation of incident nonfatal CVD outcomes in our study. Finally, traditional clinical risk factors are inexpensive, standardized, and routinely available, whereas clinical implementation of DNAm-based biomarkers would require assay standardization and cost evaluation.

Overall, DNAmScore captured residual mortality-related risk beyond PREVENT and provided modest but significant improvements when added to the clinical model. Though modest, the consistency of these improvements across subgroups and time horizons supports further study of DNAm-based biomarkers for clinical risk prediction. Additionally, because the NHANES DNAm subgroup is composed of mostly older adults, traditional risk factors may be heavy drivers of mortality in this group. As DNAm profiling and multi-omics testing become more scalable, future studies should emphasize external validation, younger cohorts, incident CVD outcomes, and development of CVD-specific DNAm risk scores compared to clinical risk equations.

## Acknowledgements

The authors thank the National Center for Health Statistics, the Centers for Disease Control, and the NHANES participants and staff for making these data publicly available.

## Data Availability Statement

All data used in this study are publicly available from the National Health and Nutrition Examination Survey (NHANES) and the public-use linked mortality files from the National Center for Health Statistics. Code is available from the corresponding author upon request.

## Ethics Statement

This study used publicly available, deidentified NHANES data and public-use linked mortality files. The original NHANES protocols were approved by the National Center for Health Statistics Research Ethics Review Board, and all participants provided informed consent. Because this secondary analysis used deidentified public-use data, additional institutional review board approval was not required.

## Author Contributions

D.G.X. and M.A.N.B. conceived and designed the study. D.G.X. performed the statistical analyses and drafted the manuscript. M.A.N.B. supervised the study. All authors contributed to interpretation of the results, revised the manuscript critically for important intellectual content, and approved the final version.

